# Blood-based host biomarker diagnostics in active case finding for pulmonary tuberculosis: a diagnostic case-control study

**DOI:** 10.1101/2020.12.27.20248917

**Authors:** Flora Martinez Figueira Moreira, Renu Verma, Paulo Cesar Pereira dos Santos, Alessandra Leite, Andrea da Silva Santos, Rafaele Carla Pivetta de Araujo, Bruna Oliveira da Silva, Júlio Henrique Ferreira de Sá Queiroz, David H. Persing, Erik Södersten, Devasena Gnanashanmugam, Purvesh Khatri, Julio Croda, Jason R. Andrews

**Affiliations:** Faculty of Health Sciences, Federal University of Grande Dourados, Dourados, MS, Brazil; Stanford University School of Medicine, Stanford, CA, USA; Cepheid, Sunnyvale, CA, USA; Cepheid AB, Solna, Sweden; Oswaldo Cruz Foundation, Campo Grande, MS, Brazil; School of Medicine, Federal University of Mato Grosso do Sul, Campo Grande, MS, Brazil; Yale School of Public Health, New Haven, CT, USA

**Keywords:** tuberculosis, diagnostic, host response, biomarker, triage, active case finding

## Abstract

**Background:** There is a need to identify scalable tuberculosis screening strategies among high burden populations. The WHO has identified a non-sputum-based triage test as a development priority.

**Methods:** We performed a diagnostic case-control study of point-of-care C-reactive protein (CRP) and Xpert-MTB-Host-Response (Xpert-MTB-HR) assays in the context of a mass screening program for tuberculosis in two prisons in Brazil. All incarcerated individuals irrespective of symptoms were screened by sputum Xpert-MTB/RIF and sputum culture. Among consecutive, Xpert-MTB/RIF or culture-confirmed cases and Xpert-MTB/RIF and culture-negative controls, CRP was quantified in serum by a point-of-care assay (iChroma-II) and a 3-gene expression score was quantified from whole blood using the Xpert-MTB-HR cartridge. We evaluated receiver operating characteristic area under the curve (AUC) and assessed specificity at 90% sensitivity and sensitivity at 70% specificity, consistent with WHO target product profile (TPP) benchmarks.

**Findings:** Two hundred controls and 100 culture- or Xpert-positive tuberculosis cases were included. Half of tuberculosis cases and 11% of controls reported any tuberculosis symptoms. AUC for CRP was 0·79 (95% CI: 0·73-0·84) and for Xpert-MTB-HR was 0·84 (95% CI: 0·79-0·89). At 90% sensitivity, Xpert-MTB-HR had significantly higher specificity (53·0%, 95% CI: 45·0-69·0%) than CRP (28·1%, 95% CI: 20·2-41·8%) (p=0·003), both well below the TPP benchmark of 70%. Among individuals with medium or high sputum Xpert semi-quantitative load, sensitivity (at 70% specificity) of CRP (90·3%, 95% CI: 74·2-98·0) and Xpert-MTB-HR (96·8%, 95% CI: 83·3-99·9%) was higher.

**Interpretation:** For active case finding in this high tuberculosis-burden setting, CRP and Xpert-MTB-HR did not meet TPP benchmarks for a triage test. However, Xpert-MTB-HR was highly sensitive in detecting individuals with medium or high sputum bacillary burden.

**Funding:** National Institutes of Health (R01 AI130058 and R01 AI149620) and Brazilian National Council for Scientific and Technological Development (CNPq-404182/2019-4).

## Introduction

Despite advances in its diagnosis and treatment, tuberculosis (TB) is still the leading cause of death among infectious diseases worldwide. It is estimated that 10 million people developed the disease and 1·4 million died from the infection in 2019.^1^ Delays in diagnosis of active TB contribute to ongoing transmission of infection and hinder the reduction of disease burden. The World Health Organization’s “End TB Strategy” proposed a set of actions to reduce the global incidence of TB by 2035, among which are the development of new efficient clinical tools for a fast and accurate diagnosis and systematic screening in high-risk groups to contribute to early detection of the diseases.^2–4^

Timely detection and treatment of cases are central elements in the control of TB transmission. This is particularly critical in high TB risk environments, such as prisons. Globally, prisons consistently harbor a very high burden of TB, with rates that are often >30 times higher than those of local non-incarcerated communities.^5^ Overcrowding, insufficient sanitation, poor ventilation and inadequate health care in prisons facilitate the spread and persistence of TB and other infectious diseases in these environments. In Central and South America, TB notifications in prisons have increased by 269% since 2000, driven primarily by growth in the size of the incarcerated population.^6^ Recent studies in Brazil have demonstrated that the prevalence of active TB among incarcerated individuals exceeds 3 900 cases per 100 000 persons, among the highest reported in the world.^7^ There is an urgent need for more effective strategies for early case detection among high TB burden populations.

One of major obstacles to early TB diagnosis is that most approaches depend upon testing of sputum, which individuals with less advanced disease may not consistently produce. In response to this challenge, the WHO has endorsed a target product profile (TPP) for a non-sputum based triage test that achieves high sensitivity in screening for active TB.^3^ There has been considerable effort undertaken to develop and validate blood-based biomarkers for TB, many focusing upon host immune response markers, including metabolomic, proteomic and transcriptomic signatures of disease.^8^ Sweeney and colleagues found that a score based on three differentially expressed genes (*GBP5* [Guanylate Binding Protein 5], *DUSP3* [Dual Specificity Protein Phosphatase 3] and *KLF2* [Kruppel-like factor 2]) had high accuracy as a screening test for active TB across multiple published cohorts. ^9^ This signature was subsequently validated for diagnosis of TB and prediction of progression from infection to disease in independent cohorts.^10^ Based on these findings, Cepheid (Sunnyvale, CA, EUA) developed an assay on their Xpert platform, cartridge-based RT-PCR assay (Xpert-MTB-HR) that quantifies the relative mRNA levels of these 3 genes in a whole blood sample. A recent study demonstrated its potential in triage and diagnosis of active TB among people living with HIV and presenting to healthcare facilities with TB symptoms.^11^ Whether this assay would perform well for active case finding, in which individuals are proactively screened for tuberculosis irrespective of symptoms, in a low HIV prevalence setting is unknown. To address this knowledge gap, we performed a diagnostic case-control study nested within a prospective active TB case finding study in prisons, evaluating the accuracy of a blood based 3-gene signature assay as potential in triage and diagnosis of active TB.

## Methods

### Study design and participants

This nested diagnostic case-control study was part of larger prospective mass TB screening study conducted in two male prisons (Estabelecimento Penal Jair Ferreira de Carvalho [EPJFC] and Penitenciaria Estadual de Dourados [PED] in Mato Grosso do Sul, Brazil.^7^ Between April 2018 and May 2020, we recruited all incarcerated individuals ages ≥18 years at both study prisons for TB screening irrespective of symptoms. After obtaining informed consent, we administered a standardized questionnaire to collect demographic and clinical information, including TB related symptoms as defined by WHO guidelines.^12^ The selection criteria for this study were the collection one spot sputum sample at enrollment and availability of a whole-blood sample. For this nested case-control study, we included consecutive participants who met case and control definitions (as described below) and had serum and PAXgene-preserved whole blood available. We did not exclude individuals with HIV or prior tuberculosis. Cases and controls were selected based on meeting outcome definitions and prior to performing the diagnostic assays under investigation in this study.

### Laboratory procedures

All participants were instructed to produce a sputum sample with a target volume of at least 2 mL. Sputum samples were tested at study entry by Xpert MTB/RIF G4 (prior to January, 2020) or Xpert Ultra (after January, 2020; Cepheid, Sunnyvale, CA, USA) along with culture on Ogawa-Kudoh media. *M. tuberculosis* was confirmed from culture growth by an immunochromatographic assay (TB Ag MPT64 Rapid Test, Standard Diagnostics, Seoul, South Korea). Upon diagnosis of a case by positive sputum Xpert or culture, we collected venous blood samples in PAXgene RNA tubes (Becton Dickinson, Franklin Lakes, USA) to preserve RNA and in BD Vacutainer tubes (Becton Dickinson, Franklin Lakes, USA) to obtain serum. We similarly collected blood from consecutive, Xpert-negative participants as potential controls and performed QuantiFERON-TB Gold Plus assays (Qiagen, Hilden, Germany) according to the manufacturer’s instructions. We stopped control recruitment upon identifying 200 Xpert negative individuals with valid QuantiFERON results. PAXgene tubes and serum samples were all collected within 14 days of first sputum sample collection (92% on same day) and were preserved in temperature-controlled freezers at −80° C from collection until testing. C-reactive protein (CRP) concentrations were measured on serum samples with a point-of-care assay (iChroma™ II, Boditech, South Korea), which has a dynamic range of 2·5-300 mg/L. The cartridge-based RT-PCR assay (Xpert-MTB-HR) was performed using 380 uL of PAXgene-preserved whole blood. The aliquot was centrifuged, supernatant decanted, and pellet was resuspended in a lysis buffer and vortexed before transfer to the Xpert-MTB-HR cartridge. The samples then were analyzed on a GeneXpert machine, which generates a TB-score calculated from cycle threshold values for the 3-genes according to the formula developed by Sweeney and colleagues.^10^ When using PCR-based measurement technology, lower scores by this method have been associated with TB disease. Sample preparation time was 15 minutes and GeneXpert run time was 50 minutes. All participants were administered a rapid HIV test and evaluated through a nursing and medical examination.

### Outcome Definitions

We followed national Brazilian guidelines and WHO definitions for TB diagnosis. We defined a microbiologically confirmed TB case as any individual with a positive sputum Xpert MTB/RIF or sputum culture. Non-microbiologically confirmed (clinically defined) TB cases were not available for inclusion in this study. Controls were defined as individuals with negative sputum Xpert MTB/RIF and culture, who were clinically evaluated and not believed to have active TB disease. All controls had been previously screened within the past year and had a chest x-ray score by CAD4TB < 60. Additionally, we searched the state Sistema de Informação de Agravos de Notificação (SINAN) data, in which all tuberculosis cases are reported, for six months following enrollment to determine whether any of the control participants were subsequently diagnosed with TB. QFT-Plus results were interpreted as positive when IFN-γ concentration in the *M. tuberculosis* (*Mtb*) antigen tube was ≥0·35 IU/mL as defined by the manufacturer.

### Statistical analysis

We compared Xpert-MTB-HR scores and serum CRP concentrations between microbiologically confirmed TB cases and controls, using Wilcoxon sign rank tests for paired samples. We evaluated the linear correlation between Xpert-MTB-HR and CRP concentrations among cases and controls. For CRP in linear analyses, values less than 2·5 mg/L (the lower limit of the reporting range) were set as 2·5 mg/L. We also compared Xpert-MTB-HR scores and serum CRP concentrations among cases with controls stratified by QFT-Plus positivity status. We calculated the receiver operating characteristic (ROC) area under the curve (AUC) comparing TB cases and controls for Xpert-MTB-HR and CRP. We compared AUC using DeLong’s test. To evaluate performance at WHO benchmarks, we calculated specificity at 90% sensitivity, and sensitivity at 70% specificity, along with 95% confidence intervals. We further investigated the sensitivity according to Xpert MTB/RIF cycle threshold (Ct) value and semi-quantitative load from sputum samples. This analysis was restricted to cases identified by Xpert MTB/RIF G4, as semiquantitative results and cycle thresholds are different for Xpert Ultra. We examined sensitivity for Xpert MTB/RIF Ct under 28·0, a threshold identified by a recent multi-country study as being 95% sensitive and 54% specific for smear positivity.^13^ We selected a sample size of 100 cases and 200 controls to achieve precision in 95% confidence interval of < +/-6.5% for the expected sensitivity of 90% and specificity of 70%. All analyses were performed using R, and ROC analyses were performed using the pROC package. ^14^

### Ethical considerations

All participants in the study provided informed consent. This study was approved by Federal University of Grande Dourados, the National Committee on Research Ethics (#44997115.1.0000.5160 and #2.195.047), and the Institutional Review Board and Stanford University (#40285). This study conforms to the Standards for Reporting of Diagnostic Accuracy Studies (STARD) reporting guideline.^15^

## Results

Between April, 2018 and May, 2020, we screened 4,250 incarcerated individuals in two prisons, of whom 1,751 were able to provide sputum for testing. Among these, 131 were positive by sputum Xpert or culture, 1456 were negative by sputum Xpert and culture, and 129 were not positive but lacked one of the two assays due to insufficient sputum volume (**Figure 1)**. From these, we selected consecutive patients for whom baseline serum and PAXgene RNA samples were available and included 100 individuals with microbiologically confirmed TB and 200 individuals who were negative for TB by sputum Xpert MTB/RIF, culture, and clinical evaluation and had valid QuantiFERON results.

**Figure 1.**
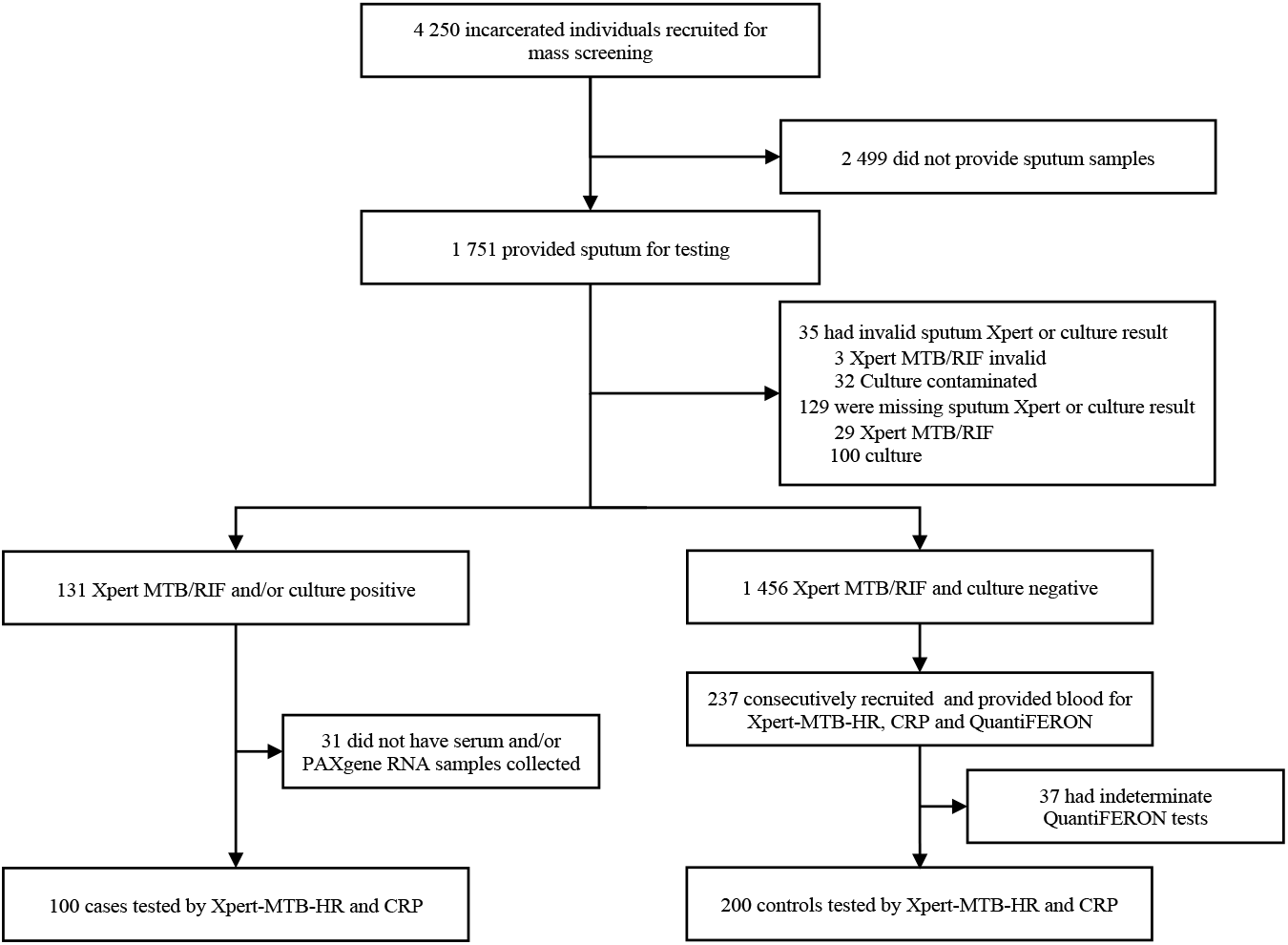
Flow diagram of study participants in mass screening and selection for diagnostic accuracy sub-study.

Among the 100 TB cases, Xpert MTB/RIF results were available for 96, and 93 (97%) were positive (85 by Xpert G4 [before January, 2020]; 8 by Xpert Ultra [after January, 2020]). Seven cases were identified by culture only (four lacked Xpert results and three were Xpert negative). All participants were men who were incarcerated at the time of enrollment, and the median age was slightly younger among TB cases (median age: 31 years, IQR 25, 37) compared with controls (median age: 34 years, IQR 28, 39). A lower proportion of cases self-identified as Indigenous (7% vs 17%; p=0·028). TB cases were more likely to report illicit drug use in the past year (80% vs 50%, p<0·0001), previous incarceration (87 vs 54%, p<0·0001), and a known TB contact (82% vs 53%, p<0·0001). Overall, half of TB cases reported any WHO TB symptom, compared with only 11% of controls (p<0·0001). The most common symptom was cough, reported by 45% of cases and 10% of controls. Four individuals with active TB (4%) were HIV co-infected, compared with none of the controls. Among controls, 92 (46%) were QFT-Plus positive (**Table 1**). Ninety-two percent (276/300) of serum and PAXgene RNA samples were collected on the same day as the first sputum sample collection and 98% (293/300) were collected within 7 days.

**Table 1.**
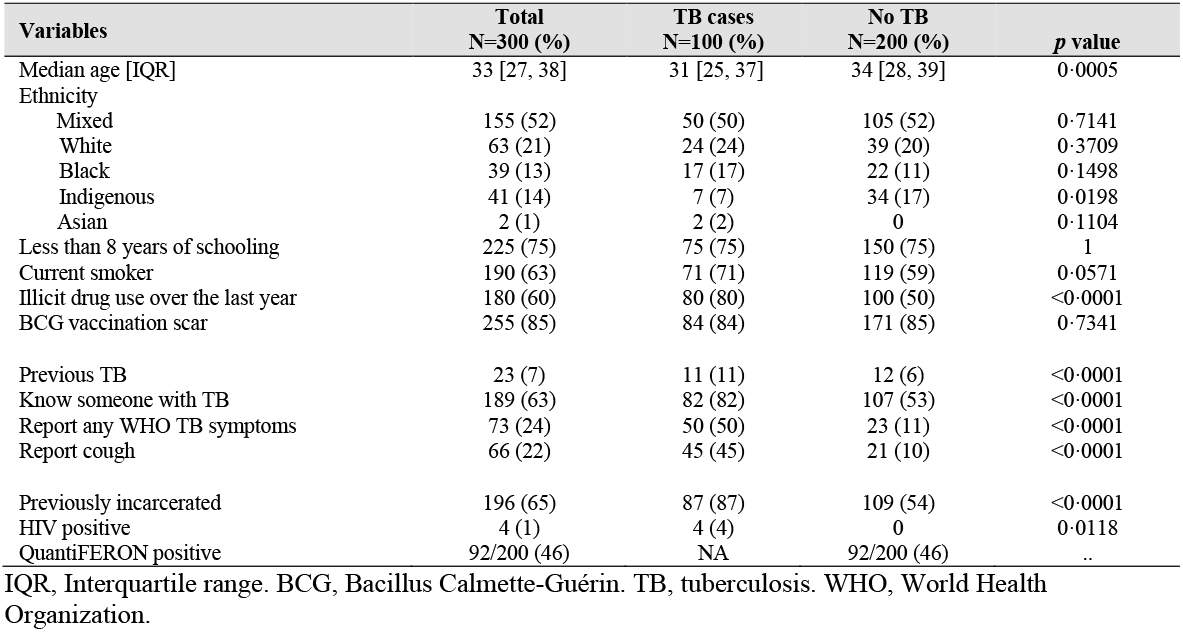
Sociodemographic and clinical characteristics of incarcerated individuals with and without TB.

| Variables | Total<br>N=300 (%) | TB cases<br>N=100 (%) | No TB<br>N=200 (%) | p value |
| --- | --- | --- | --- | --- |
| Median age [IQR] | 33 [27, 38] | 31 [25, 37] | 34 [28, 39] | 0.0005 |
| Ethnicity |  |  |  |  |
| Mixed | 155 (52) | 50 (50) | 105 (52) | 0.7141 |
| White | 63 (21) | 24 (24) | 39 (20) | 0.3709 |
| Black | 39 (13) | 17 (17) | 22 (11) | 0.1498 |
| Indigenous | 41 (14) | 7 (7) | 34 (17) | 0.0198 |
| Asian | 2 (1) | 2 (2) | 0 | 0.1104 |
| Less than 8 years of schooling | 225 (75) | 75 (75) | 150 (75) | 1 |
| Current smoker | 190 (63) | 71 (71) | 119 (59) | 0.0571 |
| Illicit drug use over the last year | 180 (60) | 80 (80) | 100 (50) | <0.0001 |
| BCG vaccination scar | 255 (85) | 84 (84) | 171 (85) | 0.7341 |
| Previous TB | 23 (7) | 11 (11) | 12 (6) | <0.0001 |
| Know someone with TB | 189 (63) | 82 (82) | 107 (53) | <0.0001 |
| Report any WHO TB symptoms | 73 (24) | 50 (50) | 23 (11) | <0.0001 |
| Report cough | 66 (22) | 45 (45) | 21 (10) | <0.0001 |
| Previously incarcerated | 196 (65) | 87 (87) | 109 (54) | <0.0001 |
| HIV positive | 4 (1) | 4 (4) | 0 | 0.0118 |
| QuantiFERON positive | 92/200 (46) | NA | 92/200 (46) | .. |

IQR, Interquartile range. BCG, Bacillus Calmette-Guérin. TB, tuberculosis. WHO, World Health Organization.

Compared with controls, TB cases had higher CRP values (9·39 vs <2·5 mg/L; p<0·0001) and lower Xpert-MTB-HR scores (1·69 vs 2·72; p<0·0001) (**Figure 2**). CRP and Xpert-MTB-HR values were negatively correlated (spearman’s rho = −0·44, p<0·0001), and the correlation was much stronger among TB cases (rho= −0·68, p<0·0001) than controls (rho = 0·01, p=0·874) (**Figure 3**). Area under the curve in distinguishing TB cases from controls was non-significantly higher for Xpert-MTB-HR (0·84, 95% CI: 0·79-0·89) than for CRP (AUC 0·79, 95% CI: 0·73-0·84; p-value for ROC comparison = 0·105) (**Figure 4**). At 90% sensitivity, the specificity of CRP was 28·1% (95% CI: 20·2-41·8%) and the specificity of Xpert-MTB-HR was 53·0% (33·5-67·0%) (p-value for comparison, 0·003). At 70% specificity, the sensitivity of CRP was 73% (95% CI: 62·1%-82·3%) and sensitivity of Xpert-MTB-HR was 81% (95% CI: 72·0-89·0%) (p-value for comparison, 0·074) **(Appendix Figure A1)**.

**Figure 2.**
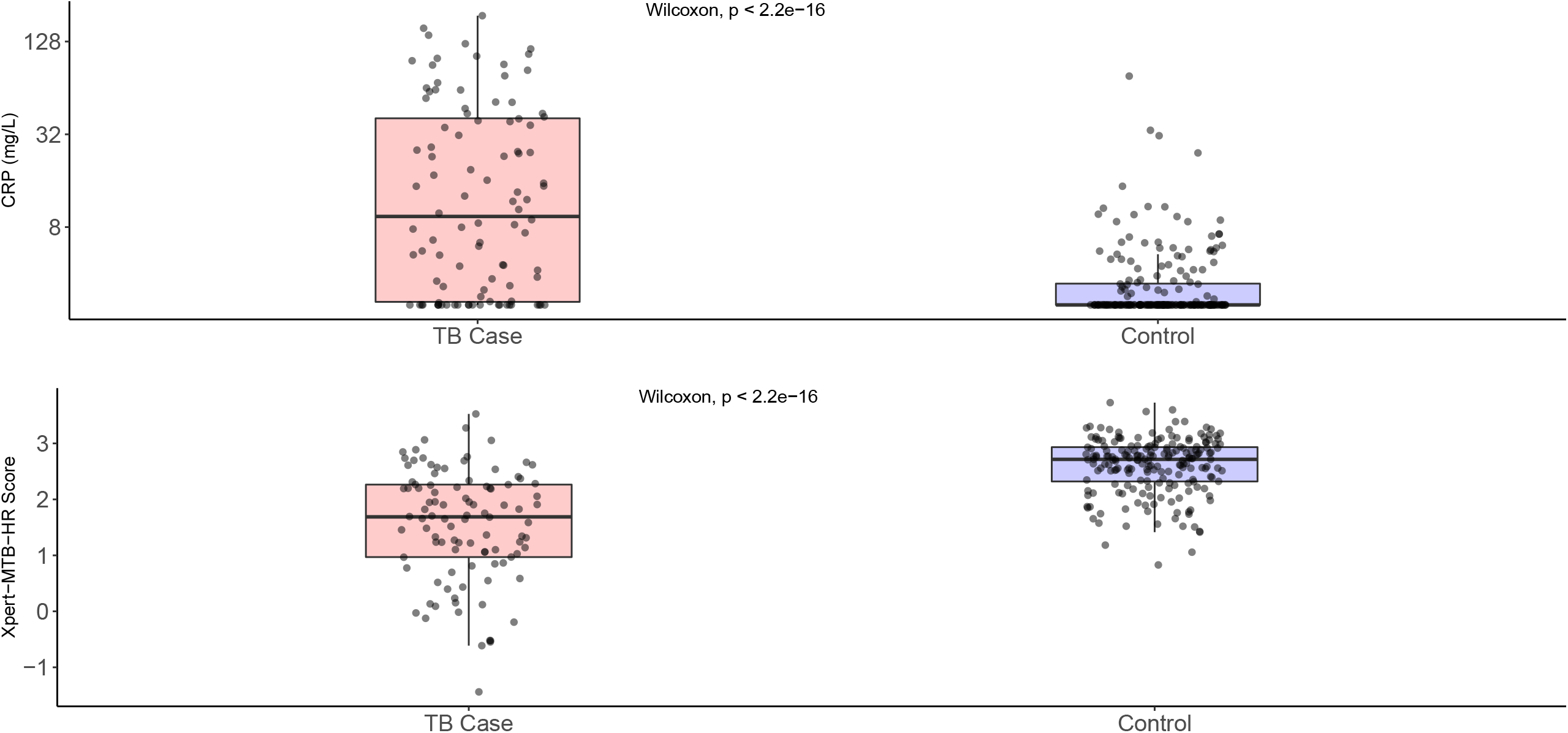
Distribution of C-reactive protein concentrations and Xpert MTB-HR scores among TB cases and controls. Box-plots comparing the distributions of C-reactive protein (top) and Xpert MTB-HR scores (bottom) between TB cases and control. CRP: C-reactive protein.

**Figure 3.**
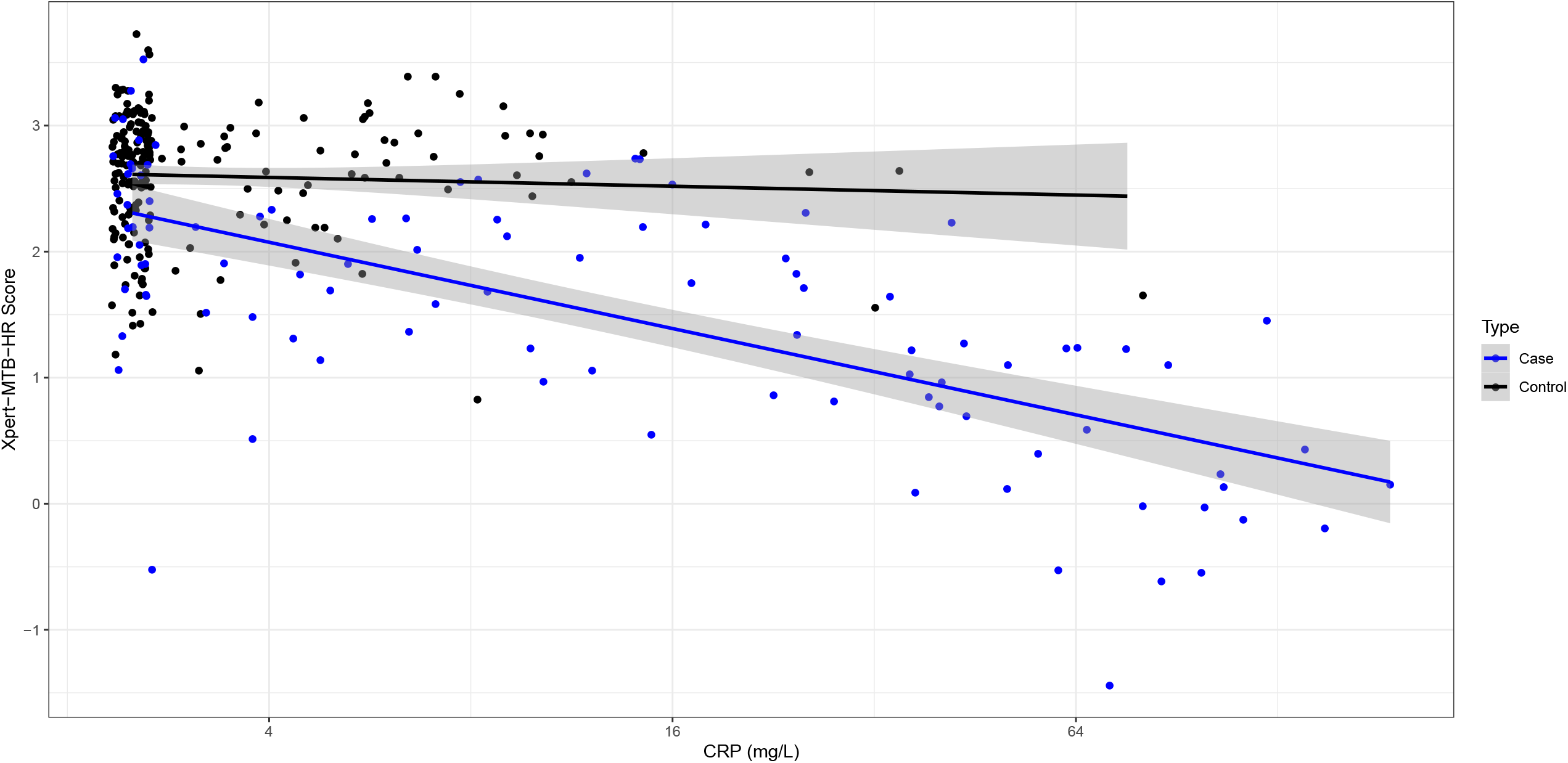
Correlation between C-reactive protein concentration and Xpert-MTB-HR Score among TB cases and controls, with fitted linear regression lines. Each dot indicates a patient observation. Black line indicates linear regression for controls and blue line indicates linear regression for cases, with shading indicating 95% confidence intervals. CRP: C-reactive protein.

**Figure 4.**
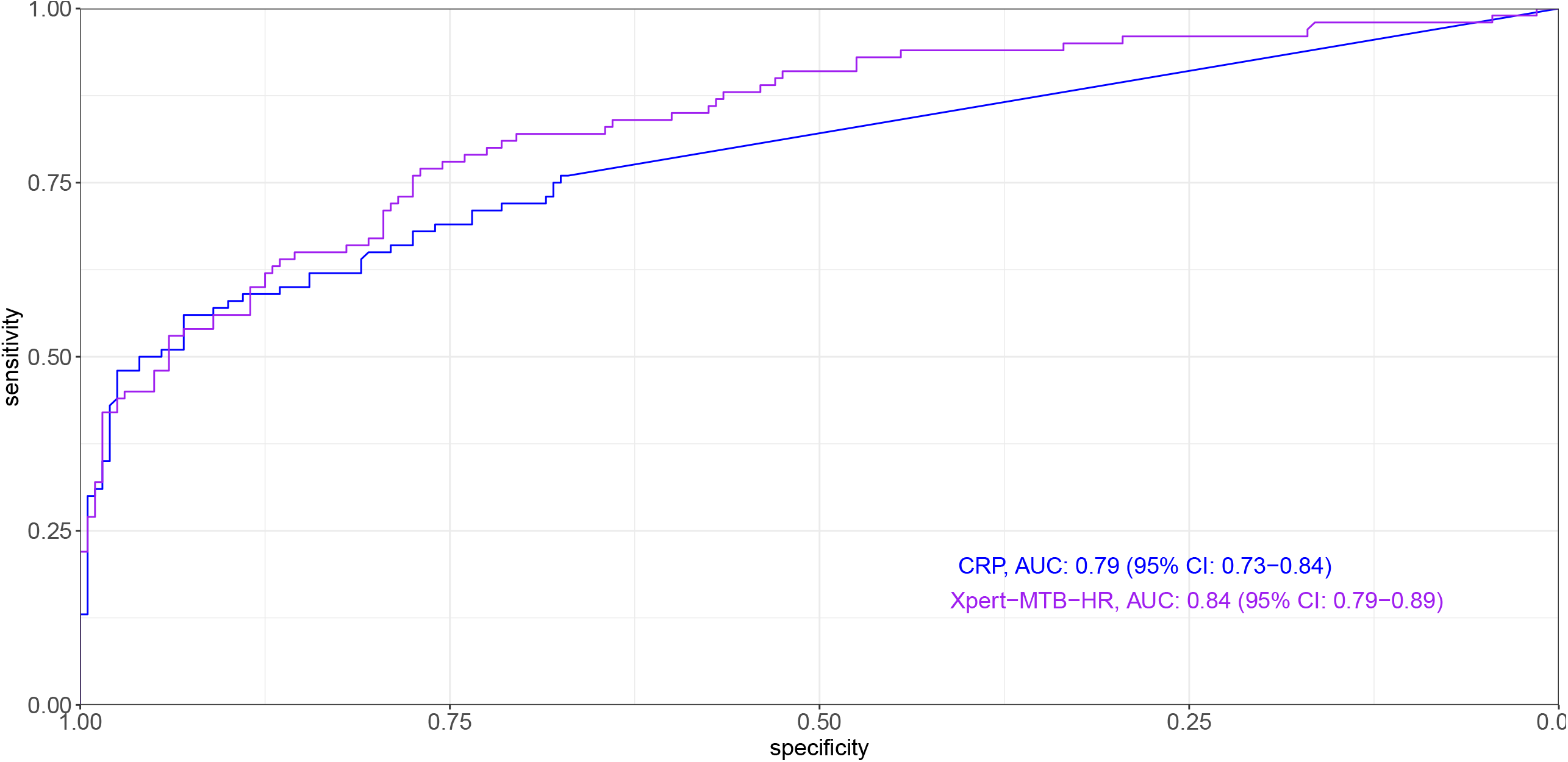
Receiver operating characteristic curves for point-of-care C-reactive protein and the Xpert MTB-HR assay. AUC: Area under the curve. CRP: C-reactive protein.

Among TB cases, CRP and Xpert-MTB-HR values correlated with sputum semi-quantitative Xpert MTB/RIF G4 values, which are based on PCR cycle thresholds. Compared with TB cases with “very low” MTB quantities, those with “medium” or “high” quantities had higher CRP (median: 20·4 vs <2·5 mg/L; p<0·0001) and lower Xpert-MTB-HR scores (median: 1·06 vs 2·20; p<0·0001) (**Figure 5a**). At the WHO TPP specificity target of 70%, the sensitivities of CRP and Xpert-MTB-HR were strongly correlated with sputum semi-quantitative Xpert MTB/RIF values (**Table 2**). Among 29 individuals with negative or very low semi-quantitative loads, CRP sensitivity was 44·8% and Xpert-MTB-HR sensitivity was 69·0% (p-value for comparison, 0·111), whereas among the 31 individuals with medium or high semiquantitative loads, CRP sensitivity was 90·3% and Xpert-MTB-HR sensitivity was 96·8% (p-value, 0·612). CRP (spearman’s rho, −0·57; p<0·0001) and Xpert-MTB-HR (spearman’s rho, 0·54, p<0·0001) values also correlated with sputum Xpert MTB/RIF Ct values (**Figure 5b**). Among 57 participants with sputum Xpert MTB/RIF Ct < 28·0, sensitivity of CRP (at 70% specificity) was 81·3% and sensitivity of Xpert-MTB-HR was 89·1%.

**Figure 5.**
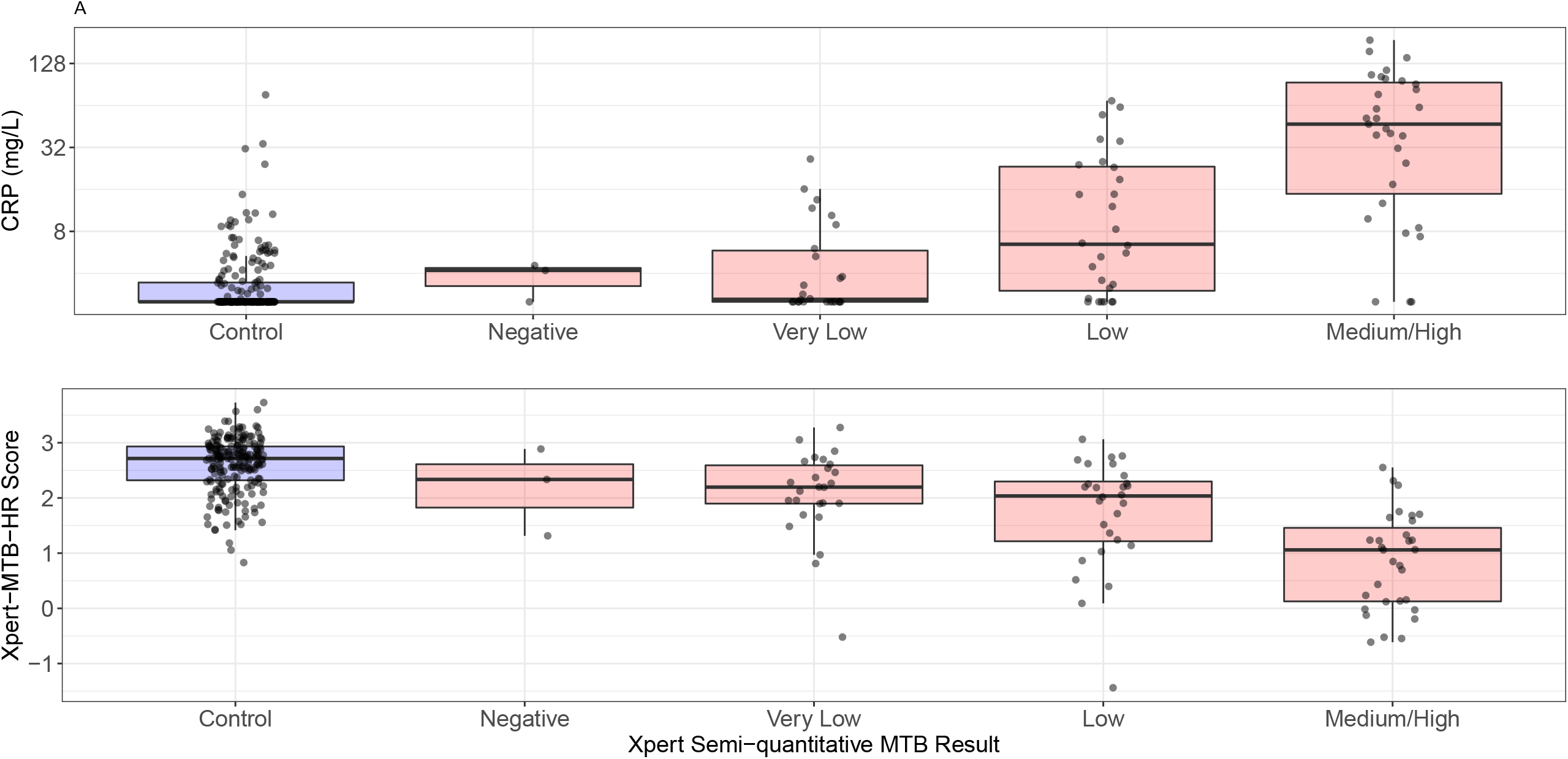
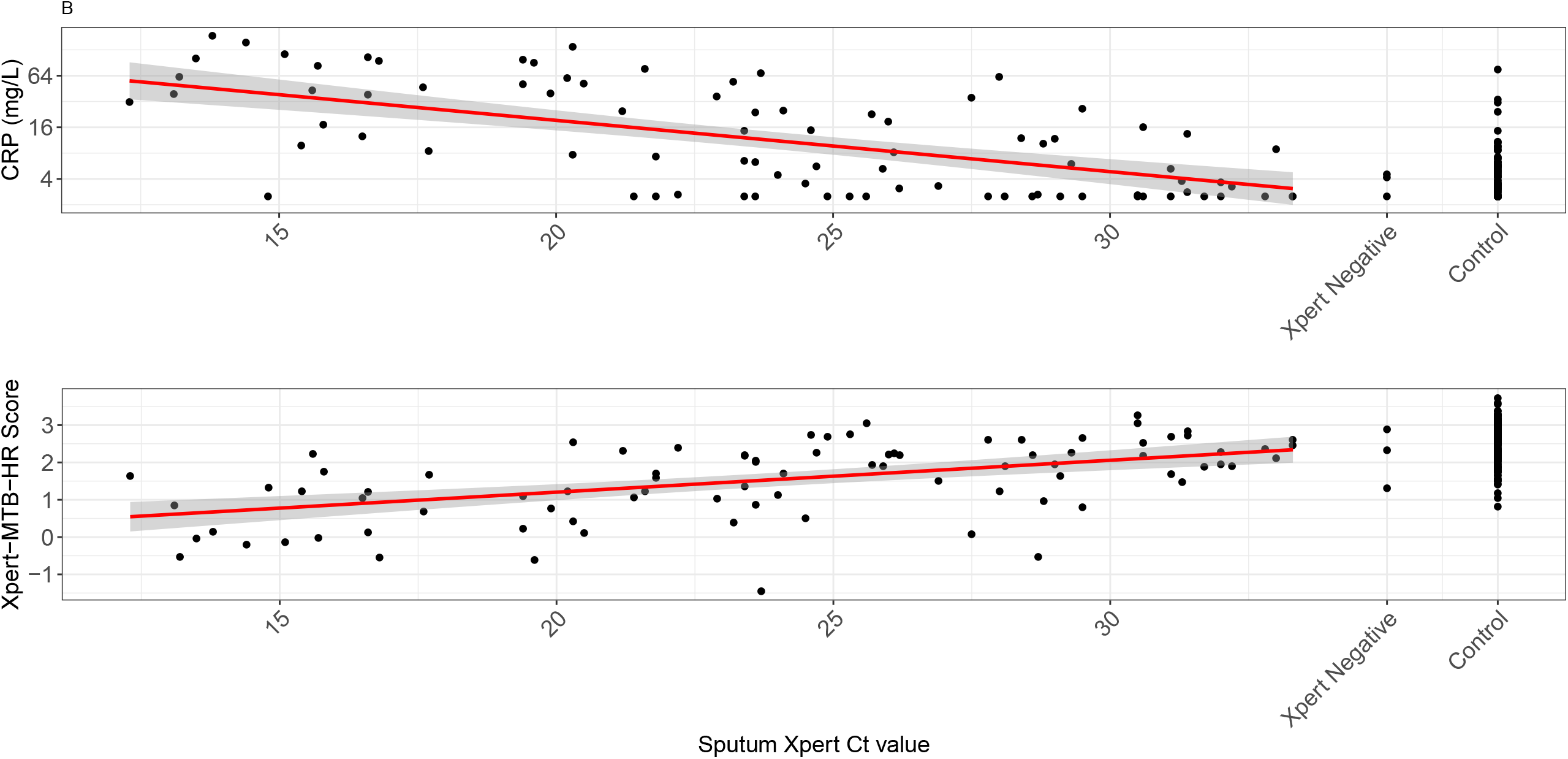
C-reactive protein and Xpert MTB-HR scores stratified by sputum Xpert semi-quantitative values (A) and cycle thresholds (B). Xpert negative, culture positive individuals (“Negative”) and Xpert- and culture-negative controls (“Control”) are depicted. CRP: C-reactive protein.

**Table 2.** Sensitivity of C-reactive protein and Xpert-MTB-HR according to sputum Xpert MTB/RIF G4 semi-quantitative load and cycle threshold values, using a threshold for specificity of 70%.

| Sputum Xpert G4 result | n | CRP |  | Xpert-MTB-HR |  |
| --- | --- | --- | --- | --- | --- |
|  |  | Positive | % Sens. (95% CI) | Positive | % Sens. (95% CI) |
| Semi-quantitative load |  |  |  |  |  |
| Negative/Very Low | 29 | 13 | 44.8 (26.4-64.3) | 20 | 69.0 (49.2-84.7) |
| Low | 28 | 21 | 75.0 (55.1-89.3) | 22 | 78.6 (59.0-91.7) |
| Medium/High | 31 | 28 | 90.3 (74.2-98.0) | 30 | 96.8 (83.3-99.9) |
| Cycle threshold |  |  |  |  |  |
| ≥28.0 | 31 | 15 | 48.4 (30.2-66.9) | 21 | 67.7 (48.6-83.3) |
| <28.0 | 57 | 47 | 82.5 (70.1-91.3) | 51 | 89.5 (78.5-96.0) |
| All confirmed cases* | 100 | 72 | 72.0 (62.1-80.5) | 82 | 82.0 (73.1-89.0) |

CRP, C-reactive protein. CI, confidence interval. \*Includes 4 culture-confirmed TB cases in whom Xpert semi-quantitative load was not available and 8 cases confirmed by Xpert Ultra.

CRP and Xpert-MTB-HR scores did not differ among TB cases with and without symptoms (p=0·541 for CRP; p=0·893 for Xpert MTB-HR). Among controls, CRP and Xpert-MTB-HR values did not differ by QFT-Plus status (p=0·211 for CRP; p=0·802 for Xpert-MTB-HR). We did not identify differences in AUC among any demographic and clinical subgroups.

## Discussion

Active case finding through systematic screening can be an effective means for early detection of TB in high-risk populations, preventing onward transmission. However, performing active case finding effectively and efficiently poses challenges, as diagnostics may not perform well in individuals who have less advanced disease, fewer symptoms, and may be unable to provide adequate sputum samples. To address this need, a target product profile was developed for a non-sputum-based triage test that could achieve high sensitivity (>90%) to identify individuals for further diagnostic evaluation.^3^ We tested two candidate triage diagnostics—point-of-care CRP and a new gene expression based rapid molecular diagnostic—from blood samples collected during systematic mass screening for TB conducted in two high TB-burden prisons, where we recently reported active TB prevalence of 3 900 per 100 000.^7^ We found that both diagnostics had moderate accuracy in distinguishing microbiologically confirmed cases from controls, with Xpert-MTB-HR achieving higher overall accuracy than CRP. At 90% sensitivity, specificity of Xpert-MTB-HR was 53%, the latter falling below the target of 70%. However, both diagnostics had greater sensitivity among individuals with higher MTB burden as measured by Xpert MTB/RIF semi-quantitative load in sputum, suggesting that they may identify individuals with greatest potential for transmission.

A number of studies have evaluated gene expression signatures for identifying individuals with active or incipient TB.^16–19^ However, there have been few studies investigating performance of these biomarkers in active case finding irrespective of symptoms, and these earlier studies used laboratory-based assays rather than rapid molecular diagnostics.^10^ The Xpert-MTB-HR test was implemented in a closed system cartridge that quantifies the relative mRNA levels of 3 genes in a whole blood sample. Our results corroborate a previous study conducted by Södersten and colleagues in a very different population and clinical setting—screening HIV-infected individuals presenting with TB symptoms to clinics in South Africa and Peru—which demonstrated feasibility of measuring the 3-gene signature on this platform.^11^ Consistent with this and earlier studies on the 3-gene signature, we found that TB cases had lower Xpert-MTB-HR scores, indicating higher expression levels (up-regulation) of *GBP5* and *DUSP3* compared with *KLF2*, compared with controls.^9–11,20^ The AUC in this study (0·84) was lower than that reported by Södersten (0·94 against sputum Xpert MTB/RIF),^11^ but consistent with a previous smaller evaluation of the 3-gene signature by laboratory-based qRT-PCR in this low HIV-prevalence, actively screened population (0.87).^10^

Earlier studies have also investigated CRP testing for distinguishing individuals with active TB from those without disease. High accuracy has been reported in some studies of HIV co-infected patients,^21,22^ though there have been fewer studies assessing screening among HIV-negative individuals, and performance has generally been poorer. A recent study among HIV-negative inpatients in Uganda found specificity of 21% at sensitivity of 91%.^23^ We are unaware of any studies assessing C-reactive protein in the context of mass screening for active case detection. We found the AUC to be 0·79 for the point-of-care CRP test in our study, consistent with the moderate accuracy previously reported in HIV-uninfected individuals. Notably, even among TB cases, the median CRP in our study was substantially lower (9·39 mg/L) than that reported among HIV-infected TB patients in a two previous studies (32·0 mg/L and 51·3 mg/L) by Yoon et al, which used the same device on serum and whole blood samples, respectively. ^21,24^ Our findings may indicate that HIV-negative individuals have less systemic inflammation in the setting of TB, perhaps consistent with the observation that HIV-infected individuals are more likely to have disseminated disease.^25^ Neither of the two diagnostics evaluated met the published TPP benchmarks for a non-sputum-based diagnostic (>90% sensitivity and >70% specificity);^3^ however, the Xpert-MTB-HR performed better (specificity 53% vs 28%). Both assays performed well in identifying individuals with medium or high *Mtb* semiquantitative load in sputum. At specificity of 70%, CRP identified 90% (28/31) and Xpert-MTB-HR identified 97% (30/31) patients with medium/high *Mtb* semiquantitative sputum Xpert® MTB/RIF result. Sputum Xpert Ct values and semi-quantitative load have been shown to correlate with sputum smear, ^13^ which in turn has been shown in multiple studies to predict infectiousness.^26,27^ It is thereby conceivable that both of these diagnostics can reliably identify individuals who have high bacillary burden and are more likely to transmit infection. Further studies are needed to investigate whether these markers indeed identify these high transmission risk individuals, and whether a triage test that identifies such individuals would be useful. Given that Xpert-MTB-HR had lower accuracy in this setting than sputum Xpert® MTB/RIF, for it to be impactful and cost-effective, it would need to either be lower cost or identify individuals with incipient, extrapulmonary or sputum-negative TB in order to be cost-effective.

This study should be interpreted within the context of several limitations. First, all participants were adult men. Our mass screening is focused in male prisons, as only two TB cases were reported in the past year in female prisons, and our prior studies in female prisons demonstrated a very low prevalence of TB in this state.^28^ The absence of female representation may have reduced the effects of confounding conditions like connective tissue disorders and pregnancy, both of which can lead to CRP elevation. Second, control participants all had chest radiographs with CAD4TB scores <60 one year prior, but this may have led to selection of controls with less lung pathology, limiting generalizability. However, >80% of all individuals in the study prisons have CAD4TB scores <60,^7^ suggesting that this group was still representative of the prison population. Third, because this was done within the context of an ongoing mass screening program, we used an older version of Xpert (G4) for the first part of the study and switched to the more sensitive Xpert Ultra when it became available at the site in January, 2020; because the Ct values are not comparable between the two cartridges, we only analyzed them for G4, which comprised 91% of the positive Xpert tests. Laboratory staff performing the biomarker diagnostics were not blinded to the reference diagnostics, introducing potential for bias, though both diagnostics have digital readouts and we retained electronic source files for the Xpert-MTB-HR performance. This was a retrospective case-control study nested within a prospective mass screening study, and it is possible that there was bias in selection of participants. Additionally, there could have been reporting biases of symptoms and characteristics by participants. Finally, we used Xpert and culture as the reference standards for classifying cases and controls. It possible that the reference assays were false positives, though they have very high specificity for tuberculosis. Some of the individuals classified as controls may had subclinical or incipient TB that was not detected by sputum Xpert MTB/RIF or culture; we performed one sputum Xpert MTB/RIF and culture on solid media. Using liquid media for culture, performing multiple Xpert tests or cultures, or use of PET-CT could improve classification. However, we note that all treated TB cases in this setting are mandatorily reported in an electronic database, which we reviewed six months after screening; none of the controls were diagnosed with TB in the six months following screening. This reduces the risk that controls were misclassified. Further, misclassification would likely bias differences in the biomarker diagnostics towards the null, meaning their accuracy would be underestimated.

There is an urgent need for accurate, rapid tools for screening individuals at high risk of tuberculosis, in order to focus further diagnostic testing and investigations. While Xpert MTB-HR outperformed CRP testing in the context of mass screening in a low HIV-prevalence population, it did not meet the WHO target product profile benchmarks for a triage test. These findings may be related to the spectrum of disease observed in this setting. Most TB cases in this population had low bacillary burden in their sputum, and half had no symptoms at the time diagnosis, suggesting that mass screening identified them early in their disease course, when they had less systemic inflammation. Among TB cases with medium or high bacillary burden, Xpert-MTB-HR achieved >96% sensitivity at 70% specificity, indicating that it may identify individuals with greater burden of infection and who might be more infectious. Further studies are needed to evaluate its performance for active case finding in other populations, and to determine whether it could be useful for averting transmission in high-risk settings.

## Data Availability

De-identified data will be made available by the corresponding author upon request.

## Authors’ contributions

FM, RV, JC and JA designed the study. FM, PCPdS, AL, AdSS, RCPdA, BOdS, JHFdSQ collected the data. FM and JA analyzed the data. ES, DG, and DHP developed the diagnostic assay and provided training for its use prior to study commencement; they played no role in assay performance for study samples and did not have access to study data. RV, DHP, ES, DG, PK and JC assisted with interpretation of the completed analyses. FM and JA wrote the first draft of the manuscript. All authors critically revised and approved the manuscript.

## Data sharing

De-identified data will be made available by the corresponding author upon request.

## Tables

**Table A1.** Specificity at fixed sensitivity values, and sensitivity at fixed specificity values for CRP and Xpert-MTB-HR.

| Sensitivity | Specificity |  | Sensitivity |  |  |
| --- | --- | --- | --- | --- | --- |
|  | CRP | Xpert-MTB-HR | Specificity | CRP | Xpert-MTB-HR |
| 0.70 | 0.74 | 0.80 | 0.70 | 0.73 | 0.81 |
| 0.75 | 0.67 | 0.78 | 0.75 | 0.69 | 0.78 |
| 0.80 | 0.56 | 0.73 | 0.80 | 0.65 | 0.70 |
| 0.85 | 0.42 | 0.62 | 0.85 | 0.61 | 0.64 |
| 0.90 | 0.28 | 0.53 | 0.90 | 0.58 | 0.57 |
| 0.95 | 0.14 | 0.36 | 0.95 | 0.51 | 0.48 |

CRP: C-reactive protein. Values were obtained from 5,000 bootstrap replicates.

